# Multimorbidity within households and health and social care utilisation and cost: retrospective cohort study using administrative data

**DOI:** 10.1101/2020.03.20.20022335

**Authors:** Mai Stafford, Sarah R. Deeny, Kathryn Dreyer, Jenny Shand

**Affiliations:** The Health Foundation, 8 Salisbury Square, London, EC4Y 8AP; UCLPartners, 170 Tottenham Court Road, London, W1T 7HA; University College London, 1-19 Torrington Place, London, WC1E 7HB; Care City, Maritime House, 1 Linton Road, Barking, IG11 8HG

**Keywords:** Multiple conditions, linked data, electronic health records

## Abstract

**Background:** The daily management of long-term conditions falls primarily on individuals and their informal carers, but the household context and its impact on health and social care activity among people with multimorbidity is understudied.

**Methods:** Linked data from health providers and local government in Barking and Dagenham provided a retrospective cohort of people aged 50+ in two-person households between April 2016 and March 2018. Two-part regression models were applied to estimate annualised use and cost of hospital, primary, community, mental health and social care by multimorbidity status of individuals and co-residents, adjusted for age, gender and deprivation. Applicability at the national level was tested using the Clinical Practice Research Datalink.

**Results:** Over 45% of multimorbid people in two-person households were co-resident with another multimorbid person. They were 1.14 (95% CI 1.00, 1.30) times as likely to have any community care activity and 1.24 (95% CI 0.99,1.54) times as likely to have any mental health care activity compared to those co-resident with a healthy person. They had more primary care visits (8.5 (95% CI 8.2,8.8) vs 7.9 (95% CI 7.7,8.2)) and higher primary care costs. Outpatient care and elective admissions did not differ between these groups. Findings in the national data were similar.

**Conclusions:** Care utilisation for people with multimorbidity varies by household context. There may be potential for connecting health and other community service input across household members.

## Introduction

As the population ages and the number of people with multiple long-term conditions (multimorbidity) grows, meeting their needs will be one of the biggest challenges facing health and social care systems. Recent UK data shows that 23-27% of people have two or more conditions and their care needs account for over 50% of primary and secondary care costs and a substantial portion of community and social care costs.[1] [2] [3] Trials of initiatives to improve outcomes and reduce emergency secondary care use of those with multimorbidity have not shown immediate success.[4] [5] Therefore, more is needed to understand the drivers of care use and costs for those living with multimorbidity, and identify ways their care could be improved.

Commonly, the daily responsibility for managing and coordinating complex care for their conditions falls primarily on the individuals themselves and on their informal carers. Around half of carers in England provide care for someone in the same household.[6] However, studies of the household context and its impact on the use and cost of services among people with multimorbidity are few in number and have focused exclusively on household size.[7] The impact of household members’ health status on health and social care use has not been studied. Co-residents are themselves at increased risk of having long-term conditions because of shared lifestyle and social risk factors and, in the case of spouses/partners, the tendency to select a similar partner. Concordance within couples and households has been shown for psychiatric and medical conditions, pain, and health behaviours [8] [9] [10] though not yet for multimorbidity. Poorer social support is one explanation for the greater health care utilisation of those living alone.[11] Available support might also be lower in households with multiple people with multimorbidity since illness of other household members may make it more difficult for them to provide practical, financial or emotional support.

The aim of the current study was to test the hypothesis that, conditional on household size, being in a household with another person with multimorbidity is associated with higher utilisation and cost of primary, community and secondary health care and formal social care among people with multimorbidity. Difficulty in identifying households within electronic health records (EHRs) likely contributes to the lack of research in this area. To address this, we used a local sample of EHRs linked to detailed household composition data from local authority records and replicated the analysis in a national study where co-residence was inferred from anonymised address data.

## Methods

The study focused on people aged 50 and over living in two-person households. We excluded single-person households because our aim was to test the effect of co-resident’s health status on care use and costs. We excluded people in households with three or more occupants to exclude institutions and because of the challenge of interpreting results where there could be multiple people providing help.

### Primary analytical sample - Barking & Dagenham

This individual-level dataset links information from local government services, health providers and health commissioners in the area. It includes sociodemographic, health and household information alongside activity data for five settings of care (primary care, hospital, community, inpatient and outpatient mental health services, and social care).

Confirmed residents of Barking and Dagenham from 1^st^ April 2016 to 31^st^ March 2018 were included. Those who moved out of Barking and Dagenham or who died before the 1st April 2018 were excluded, given the known increase in health care utilisation at the end of life[12] that could bias results. Individuals were grouped into households using the unique property reference number in local government records. The analytical sample was 9,222 individuals in 4,611 two-person households.

### Measuring multimorbidity

We included long-term conditions that lead to a significant need for treatment or have been linked to poorer quality of life or greater risk of premature death.[1] [13] [14] Sixteen long-term mental and physical health conditions were identified from diagnosis codes recorded in primary care (Supplementary Table 1). People with 2+ conditions were classified as “multimorbid” and those with 0-1 condition as “healthy”. We then categorised each person into one of four household multimorbidity categories: multimorbid person co-resident with healthy person (“MM/healthy” - the reference group); multimorbid person co-resident with another multimorbid person (“MM/MM”); healthy person co-resident with healthy person (“healthy/healthy”); healthy person co-resident with multimorbid person (“healthy/MM”).

### Health and social care activity and cost

We examined health and social care activity and associated costs over two years. Outcomes included: number and cost of primary care consultations with a general practitioner (GP), nurse or other clinical staff; number of days in hospital and the cost of hospital care (also broken down into cost of outpatient consultations, cost of elective admissions, cost of non-elective admissions, and cost of emergency department attendances); cost of mental health inpatient and outpatient care; cost of community health services; and cost of local government funded social care.

Local unit level costs were used for community and mental health activity and social care services. All other costs were based on activity using national reference costs for the relevant year. Mean cost for the relevant activity was used where an activity code could not be matched to a national reference cost. Costs were indexed to 2018 prices. See Supplementary Table 2 for more detail on the costing method.

### Statistical analysis

The distributions of care costs and number of days in hospital show a substantial proportion of people having zero activity. We used two-part mixture models to account for their semi-continuous distributions. The first part used a logistic model to estimate the likelihood of having any relevant care activity or not. In other words, the first part modelled each type of care utilisation as a binary variable (any versus no care use). The second part used a gamma model to estimate the cost or utilisation among the subset where this was non-zero. Gender, age group, deprivation (Index of Multiple Deprivation quintile for the patient’s residence), and multimorbidity status were included as covariates for both parts. We also estimated the costs, number of consultations or number of days in hospital across both parts of the model combined. Analysis was done at the individual level with robust standard errors to allow for the non-independence of individuals within households.

### Replication analysis in a national sample

Nationally representative data were obtained from the Clinical Practice Research Datalink (CPRD). CPRD comprises de-identified records of over 14 million patients from consenting GP practices in the UK[15] with linkage to Hospital Episode Statistics for consenting practices in England. Individuals registered in up-to-standard practices from 1^st^ April 2014 to 31^st^ March 2016 were included.

The CPRD pseudonymised family number (based on the first line of the patient’s address) was used to select a sample where exactly two patients shared a family identifier and were registered within one year of each other. This approach excludes two-person households where members are not registered at the same GP practice. From an initial random sample of 300,000 children and adults with linked HES data, 10,528 met inclusion criteria and formed the analytical sample for this study.

The presence of 36 mental and physical health conditions (see Supplementary Table 3) recorded in primary care[16] was determined on 1^st^ April 2014 based on diagnosis (using Read codes) and prescribing data. Code lists differ from those used in the local sample but there is considerable overlap with the 16 broader groups of long-term conditions included in the primary analysis. Health care activity and cost was calculated using the same approach. Data on mental health care, community health care and social care activity and cost were not available for the national sample.

### Approval for the study

Routinely collected, retrospective, pseudonymised data were used for this analysis. Approval for secondary analysis of the national data was obtained from the Clinical Practice Research Datalink Independent Scientific Advisory Committee (protocol ISAC17_150RMn2).

## Results

### Sample description: primary sample

Multimorbidity prevalence was 43% among the over 50s living in two-person households in Barking and Dagenham. Forty-eight per cent of multimorbid people were co-resident with another multimorbid person (Table 1). Older people and people living in more deprived areas were over-represented in households with two multimorbid residents.

**Table 1.** Description of the study samples i) primary sample from Barking and Dagenham, ii) national sample from Clinical Practice Research Datalink

|  | MM / healthy co-resident |  | Both MM |  | Healthy / MM co-resident |  | Both healthy |  | Total |
| --- | --- | --- | --- | --- | --- | --- | --- | --- | --- |
|  | n | % | n | % | n | % | n | % | n |
| **B & D sample** | 2049 | 22.2 | 1884 | 20.4 | 3240 | 35.1 | 2049 | 22.2 | 9222 |
| Age |  |  |  |  |  |  |  |  |  |
| 50-59 | 337 | 14.4 | 205 | 8.8 | 1240 | 53.1 | 554 | 23.7 | 2336 |
| 60-69 | 620 | 20.9 | 566 | 19.1 | 1124 | 37.9 | 655 | 22.1 | 2,965 |
| 70-79 | 644 | 26.0 | 663 | 26.8 | 595 | 24.0 | 575 | 23.2 | 2477 |
| 80+ | 448 | 31.0 | 450 | 31.2 | 281 | 19.5 | 265 | 18.4 | 1444 |
| Gender |  |  |  |  |  |  |  |  |  |
| Male | 1044 | 22.7 | 928 | 20.1 | 1640 | 35.6 | 997 | 21.6 | 4609 |
| Female | 1005 | 21.8 | 956 | 20.7 | 1600 | 34.7 | 1052 | 22.8 | 4613 |
| IMD quintile |  |  |  |  |  |  |  |  |  |
| 1 (least deprived) | 0 | 0.0 | 0 | 0.0 | 0 | 0.0 | 0 | 0.0 | 0 |
| 2 | 0 | 0.0 | 0 | 0.0 | 0 | 0.0 | 0 | 0.0 | 0 |
| 3 | 192 | 21.7 | 163 | 18.4 | 339 | 38.3 | 192 | 21.7 | 886 |
| 4 | 768 | 22.5 | 617 | 18.1 | 1263 | 37.0 | 770 | 22.5 | 3418 |
| 5 (most deprived) | 1089 | 22.1 | 1104 | 22.4 | 1638 | 33.3 | 1087 | 22.1 | 4918 |
| Carer code present | 134 | 6.5 | 158 | 8.4 | 81 | 2.5 | 56 | 2.7 | 429 |
| **CPRD sample** | 2107 | 20.0 | 2940 | 27.9 | 3374 | 32.0 | 2107 | 20.0 | 10528 |
| Age |  |  |  |  |  |  |  |  |  |
| 50-59 | 530 | 15.8 | 372 | 11.1 | 1770 | 52.7 | 687 | 20.5 | 3359 |
| 60-69 | 791 | 21.4 | 915 | 24.7 | 1179 | 31.9 | 817 | 22.1 | 3702 |
| 70-79 | 568 | 22.8 | 1081 | 43.3 | 377 | 15.1 | 471 | 18.9 | 2497 |
| 80+ | 218 | 22.5 | 572 | 59.0 | 48 | 5.0 | 132 | 13.6 | 910 |
| Gender |  |  |  |  |  |  |  |  |  |
| Male | 1171 | 22.3 | 1464 | 27.8 | 1697 | 32.3 | 928 | 17.6 | 5260 |
| Female | 936 | 17.8 | 1476 | 28.0 | 1677 | 31.8 | 1179 | 22.4 | 5268 |
| IMD quintile |  |  |  |  |  |  |  |  |  |
| 1 (least deprived) | 728 | 20.0 | 871 | 24.0 | 1306 | 35.9 | 729 | 20.1 | 3634 |
| 2 | 491 | 19.8 | 674 | 27.1 | 825 | 33.2 | 496 | 20.0 | 2586 |
| 3 | 426 | 20.2 | 598 | 28.3 | 662 | 31.4 | 425 | 20.1 | 2111 |
| 4 | 302 | 19.9 | 510 | 33.6 | 410 | 27.0 | 296 | 19.5 | 1518 |
| 5 (most deprived) | 160 | 20.5 | 287 | 36.8 | 171 | 22.0 | 161 | 20.7 | 779 |
| Carer code present | 48 | 2.3 | 197 | 6.7 | 48 | 1.4 | 81 | 3.8 | 374 |

Over 97% of people had at least one primary care consultation during follow-up, but over 30% had no outpatient attendance, 80% had no emergency department attendance and over 80% had no inpatient admission (Supplementary Table 4).

### Model results: primary sample

Table 2 summarises the estimates from the two-part models. We first estimated the association between each person’s household multimorbidity status and the likelihood of having any care activity during the follow up time (see column labelled “OR for any activity”). We focus on differences between a multimorbid person co-resident with another multimorbid person and one co-resident with a healthy person. Controlling for gender, age, and socioeconomic deprivation, we found no statistically significant differences between these groups in the likelihood of having any activity for the care outcomes, with one exception. The “Both MM” group were 1.14 (95% CI 1.00, 1.30) times as likely to have any community care activity. In addition, the “Both MM” group were 1.24 (95% CI 0.99, 1.54) times as likely to have any mental health care activity, and 1.24 (95% CI 0.96, 1.59) times as likely to have any social care activity as the “MM/healthy” reference group.

**Table 2.** Two-part models for health and social care activity: Barking and Dagenham sample of N=9222 people age 50+ in two-person households

|  | OR for<br>any activity | 95%<br>LL | 95%<br>UL | Exp gamma<br>coefficient (in the<br>subset with any<br>activity) | 95%<br>LL | 95%<br>UL | Adjusted means<br>across both parts | 95%<br>LL | 95%<br>UL |
| --- | --- | --- | --- | --- | --- | --- | --- | --- | --- |
| Number of primary care visits |  |  |  |  |  |  |  |  |  |
| MM / healthy | Reference |  |  | Reference |  |  | 7.9 | 7.7 | 8.2 |
| Both MM | 0.44 | 0.17 | 1.11 | 1.08 | 1.03 | 1.12 | 8.5 | 8.2 | 8.8 |
| Both healthy | 0.12 | 0.06 | 0.27 | 0.62 | 0.60 | 0.65 | 4.8 | 4.7 | 4.9 |
| Healthy / MM | 0.13 | 0.06 | 0.30 | 0.66 | 0.64 | 0.69 | 5.1 | 5.0 | 5.3 |
| Primary care costs |  |  |  |  |  |  |  |  |  |
| MM / healthy | Reference |  |  | Reference |  |  | £198 | £192 | £205 |
| Both MM | 0.44 | 0.17 | 1.11 | 1.08 | 1.03 | 1.13 | £213 | £205 | £220 |
| Both healthy | 0.12 | 0.06 | 0.27 | 0.64 | 0.62 | 0.67 | £124 | £121 | £128 |
| Healthy / MM | 0.13 | 0.06 | 0.30 | 0.69 | 0.66 | 0.72 | £134 | £130 | £138 |
| Outpatient costs |  |  |  |  |  |  |  |  |  |
| MM / healthy | Reference |  |  | Reference |  |  | £312 | £294 | £330 |
| Both MM | 1.03 | 0.89 | 1.19 | 0.93 | 0.86 | 1.00 | £293 | £275 | £311 |
| Both healthy | 0.50 | 0.44 | 0.56 | 0.71 | 0.66 | 0.76 | £172 | £162 | £182 |
| Healthy / MM | 0.51 | 0.45 | 0.58 | 0.73 | 0.68 | 0.79 | £181 | £168 | £193 |
| Cost of non-elective admissions |  |  |  |  |  |  |  |  |  |
| MM / healthy | Reference |  |  | Reference |  |  | £609 | £532 | £686 |
| Both MM | 1.01 | 0.87 | 1.17 | 0.99 | 0.85 | 1.14 | £606 | £527 | £684 |
| Both healthy | 0.45 | 0.39 | 0.53 | 0.76 | 0.64 | 0.90 | £247 | £205 | £288 |
| Healthy / MM | 0.42 | 0.35 | 0.50 | 0.63 | 0.53 | 0.76 | £192 | £154 | £230 |
| Cost of elective admissions |  |  |  |  |  |  |  |  |  |
| MM / healthy | Reference |  |  | Reference |  |  | £443 | £396 | £491 |
| Both MM | 1.05 | 0.92 | 1.20 | 1.00 | 0.88 | 1.14 | £459 | £409 | £509 |
| Both healthy | 0.57 | 0.51 | 0.65 | 0.91 | 0.80 | 1.03 | £273 | £244 | £303 |
| Healthy / MM | 0.64 | 0.56 | 0.73 | 0.86 | 0.75 | 0.98 | £281 | £245 | £316 |
| ED costs |  |  |  |  |  |  |  |  |  |
| MM / healthy | Reference |  |  | Reference |  |  | £84 | £76 | £92 |
| Both MM | 1.08 | 0.95 | 1.23 | 1.04 | 0.93 | 1.16 | £91 | £82 | £99 |
| Both healthy | 0.53 | 0.47 | 0.60 | 0.76 | 0.67 | 0.85 | £43 | £38 | £47 |
| Healthy / MM | 0.52 | 0.45 | 0.59 | 0.78 | 0.68 | 0.88 | £43 | £38 | £48 |
| Total hospital costs (outpatient + admissions + ED) |  |  |  |  |  |  |  |  |  |
| MM / healthy | Reference |  |  | Reference |  |  | £1454 | £1338 | £1570 |
| Both MM | 1.06 | 0.91 | 1.24 | 0.99 | 0.89 | 1.11 | £1458 | £1338 | £1578 |
| Both healthy | 0.49 | 0.43 | 0.56 | 0.62 | 0.56 | 0.69 | £743 | £685 | £800 |
| Healthy / MM | 0.47 | 0.41 | 0.54 | 0.59 | 0.53 | 0.67 | £701 | £636 | £766 |
| Number of days in hospital |  |  |  |  |  |  |  |  |  |
| MM / healthy | Reference |  |  | Reference |  |  | 1.58 | 1.33 | 1.83 |
| Both MM | 0.97 | 0.84 | 1.12 | 0.98 | 0.80 | 1.20 | 1.53 | 1.28 | 1.77 |
| Both healthy | 0.43 | 0.37 | 0.50 | 0.80 | 0.63 | 1.00 | 0.65 | 0.51 | 0.78 |
| Healthy / MM | 0.42 | 0.35 | 0.50 | 0.67 | 0.52 | 0.87 | 0.54 | 0.41 | 0.67 |
| Community care costs |  |  |  |  |  |  |  |  |  |
| MM / healthy | Reference |  |  | Reference |  |  | £428 | £358 | £498 |
| Both MM | 1.14 | 1.00 | 1.30 | 1.20 | 0.97 | 1.49 | £559 | £468 | £650 |
| Both healthy | 0.37 | 0.32 | 0.43 | 0.69 | 0.54 | 0.89 | £141 | £111 | £171 |
| Healthy / MM | 0.36 | 0.31 | 0.42 | 0.51 | 0.39 | 0.68 | £103 | £77 | £128 |
| Mental health costs |  |  |  |  |  |  |  |  |  |
| MM / healthy | Reference |  |  | Reference |  |  | £318 | £155 | £482 |
| Both MM | 1.24 | 0.99 | 1.54 | 1.41 | 0.73 | 2.71 | £543 | £281 | £795 |
| Both healthy | 0.37 | 0.28 | 0.50 | 1.01 | 0.43 | 2.40 | £127 | £31 | £224 |
| Healthy / MM | 0.40 | 0.30 | 0.54 | 0.76 | 0.30 | 1.92 | £103 | £19 | £186 |
| Social care costs |  |  |  |  |  |  |  |  |  |
| MM / healthy | Reference |  |  | Reference |  |  | £439 | £320 | £559 |
| Both MM | 1.24 | 0.96 | 1.59 | 0.86 | 0.66 | 1.13 | £461 | £347 | £575 |
| Both healthy | 0.29 | 0.20 | 0.43 | 0.58 | 0.38 | 0.88 | £80 | £42 | £118 |
| Healthy / MM | 0.27 | 0.18 | 0.41 | 0.51 | 0.32 | 0.81 | £64 | £28 | £99 |

**Table 3.**
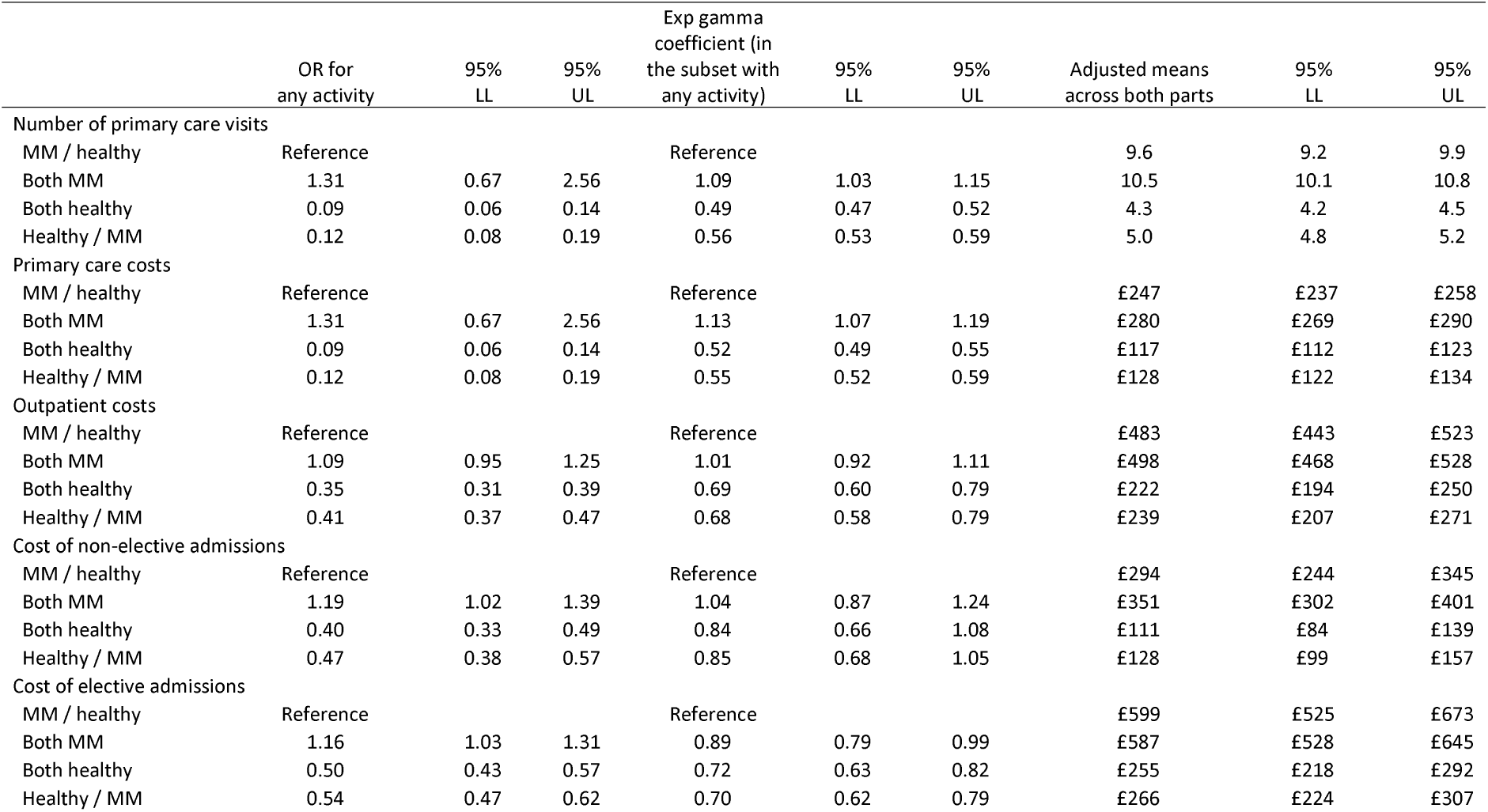
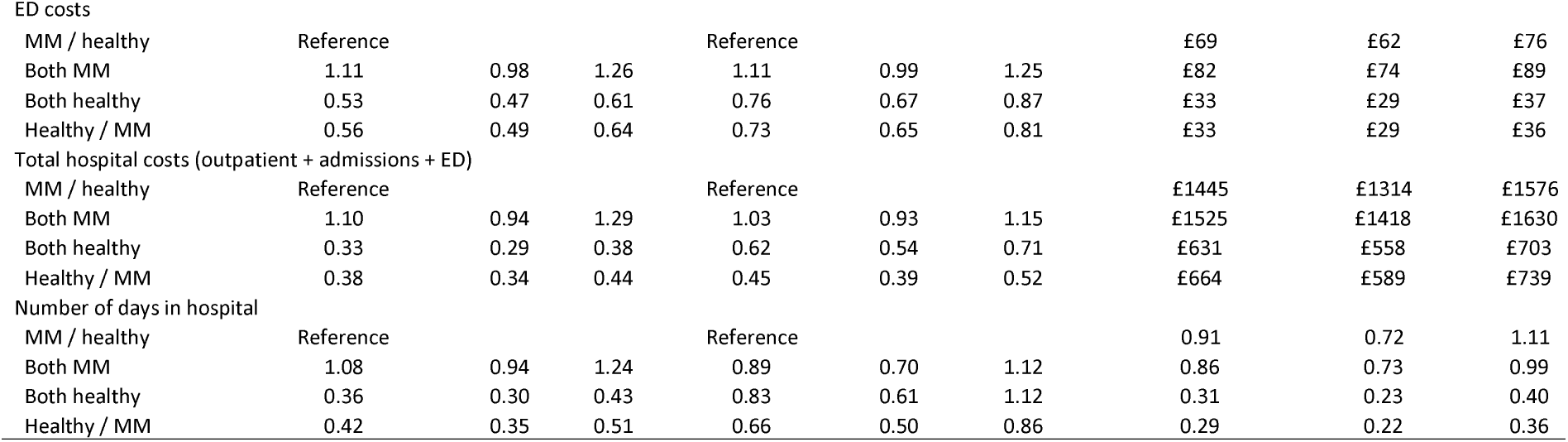
Two-part models for health care activity: CPRD sample of N=10528 people age 50+ in two-person households

The column of exponentiated gamma coefficients allows us to test the association between household multimorbidity status and level of care activity for the subset of people who have any activity. The coefficient of 1.08 (95% CI 1.03, 1.12) for the number of primary care visits shows that the adjusted difference in number of visits for a person in the “Both MM” group was 8% (95% CI 3% to 12%) higher than the number of visits for a person in the reference group. A similar difference in cost of primary care visits was also seen. Among the subsets with any activity, there was no evidence of a difference in costs between these groups for other outcomes.

The adjusted means and their confidence limits in the final three columns combine estimates from both parts of the model (i.e. including those who do and do not have the relevant activity). Adjusted mean annual primary care cost was £198 for a multimorbid person co-resident with a healthy person, £213 for a multimorbid person co-resident with another multimorbid person, and £124 where both residents were healthy. A multimorbid person co-resident with a healthy person had 7.9 primary care visits per year whereas one co-resident with another multimorbid person had 8.5 per year. Community care costs for the “Both MM” group were £559 compared with £428 for the reference group though the 95% confidence intervals for these estimates overlapped. The corresponding estimates for mental health care costs were £543 and £318 though confidence intervals for these estimates also overlapped (Figure 1).

**Figure 1.**
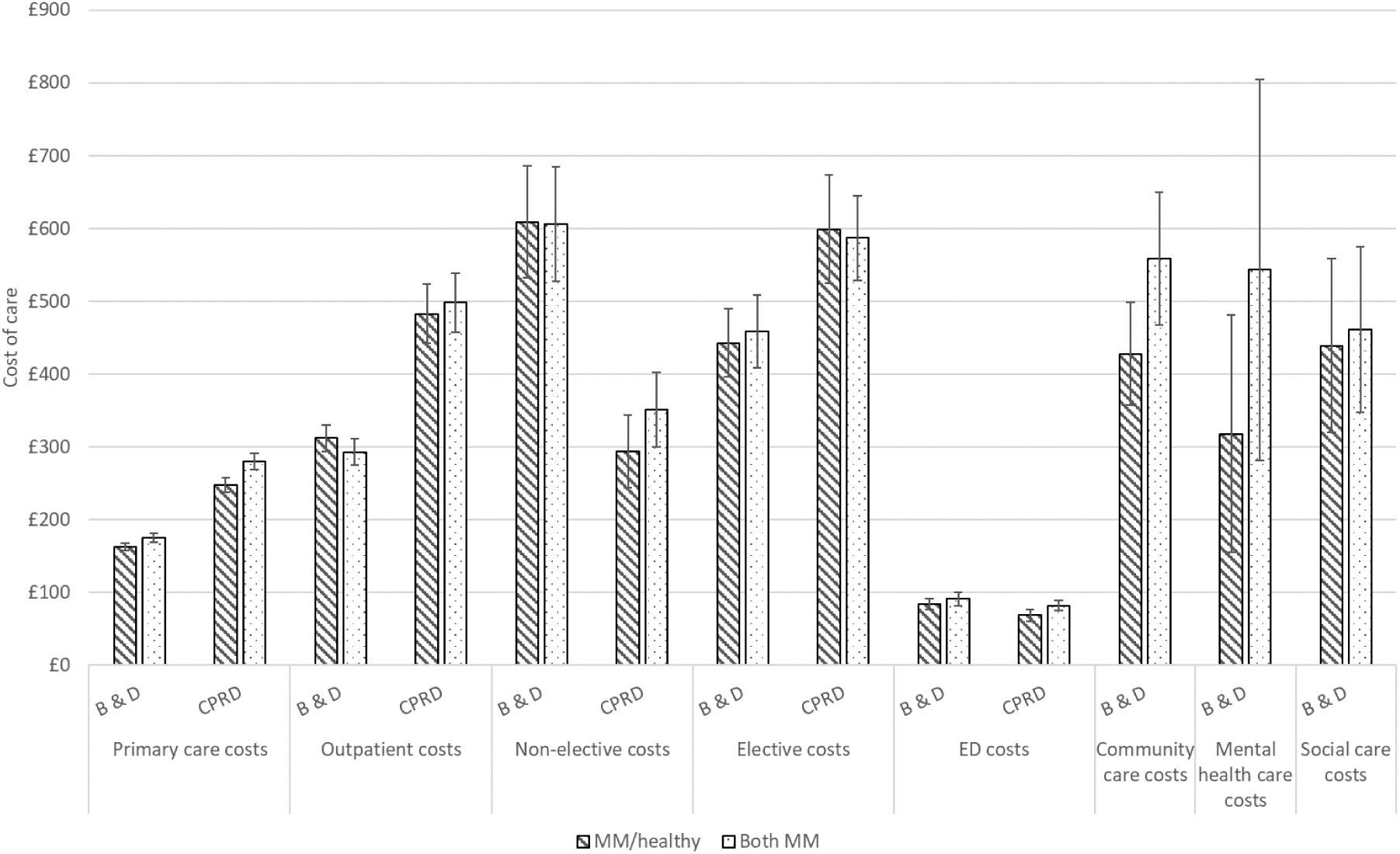
Cost of care by household multimorbidity status

Although not the focus of this study, we found that those who were not multimorbid (in either the “Both healthy” or the “Healthy/MM” group) were less likely to have any activity than the reference group. They also had lower adjusted mean costs and level of utilisation for all types of care.

### Replication in the national sample

Multimorbidity prevalence was 48% among the over 50s living in two-person households in the national sample. This is higher than in Barking and Dagenham, possibly because 36 rather than 16 conditions were counted. Fifty-eight percent of multimorbid people were co-resident with another multimorbid person and the same associations with age and deprivation were seen in the national sample.

A multimorbid person co-resident with another multimorbid person was more likely to have any non-elective hospital admission (OR 1.19 (95% 1.02,1.39)) or elective admission and more likely to have any emergency department visit (OR 1.11 (95% 0.98,1.26)) compared with the reference group.

Among the subset of patients with any primary care cost, this cost was 13% (95% CI 7%, 19%) higher in the “Both MM” group compared with the reference, and the “Both MM” group also had more primary care visits. There was a suggestion of higher emergency department costs (exponentiated gamma coefficient 1.11 (95% CI 0.99,1.25, p=0·06)) with no evidence of difference between these groups for other outcomes among those with a non-zero cost or activity.

Across both parts of the model combined, mean primary care cost was £247 for a multimorbid person co-resident with a healthy person, £280 for a multimorbid person co-resident with another multimorbid person, and £117 where both residents were healthy. The mean number of primary care consultations was 9·6 for a multimorbid person co-resident with a healthy person and 10·5 for a multimorbid person co-resident with another multimorbid person.

## Discussion

This study provides evidence that the household context matters for the care utilisation of people with multimorbidity, independently of age, gender and area deprivation. Primary care costs and number of primary care visits each year were higher for multimorbid people if they were co-resident with another multimorbid person compared with a healthy one. This evidence is based on one local and one national sample of two-person households aged fifty and over. The local sample with linked local government and health service data also indicated a greater likelihood of community care activity where a multimorbid person was co-resident with another. In the national sample, a greater likelihood of non-elective hospital activity was seen where a multimorbid person was co-resident with another.

Though this is an observational study and unobserved confounding cannot be excluded, there are three substantive mechanisms that could underlie observed associations. First, a co-resident with multimorbidity may have less capacity to provide informal support than a healthy co-resident. Lack of informal support has sometimes been associated with greater health care utilisation[17] although caregivers can also act as advocates in ways that increase some types of care use. They may help a patient overcome denial about their need for care and may be more proactive in seeking help.[18] It is not known whether this advocacy role depends on the advocate’s health status but available information about access to and benefits of health services might be greater in households where both residents have multimorbidity, resulting in greater health care use with concordant multimorbidity. Third, the multimorbid person may be providing care for their multimorbid co-resident. Providing informal care can be a stressful experience with negative health consequences, especially in the context of low levels of formal care,[19] and this could contribute to increased use of health care although carers have reported greater difficulty accessing primary care compared with non-carers[20] and may avoid treatment because of their caring responsibilities.[21] This third explanation is supported by our data showing that a healthy person co-resident with a multimorbid person also had higher primary care costs and more primary care visits than their counterparts co-resident with a healthy person.

We were unable to test these explanations because informal care-giving and receiving is not well recorded in administrative records. In this study, less than 5% had a Read code relating to caring whereas the latest census found over 17% of people aged 50 and over in England were providing informal care for someone with a long-term physical or mental health condition or disability.[22] The long-term plan for the NHS[23] and other government initiatives[24] commit to better identifying and supporting carers and our study suggests this has the potential to benefit people living with multimorbidity as both givers and recipients of informal care.

### Strengths and limitations

Two-part models were used to model cost and utilisation outcomes. Outcomes were based on electronic health records and not subject to recall or reporting bias. Two large samples were used to test our hypothesis. Similar proportions of own and co-resident multimorbidity were found in the two data sources. In the Barking and Dagenham sample, local authority data provided a detailed measure of household composition and allowed us to accurately identify two-person households. For primary care and hospital care, we were able to replicate the findings in a nationally representative sample. This provides reassurance that the pseudonymised variable based on the first line of the patient’s last known address in combination with current registration date is a reasonable approach to identifying two-person households in primary care records.

We could not distinguish partners from other types of co-resident in either sample. Marital or partnership status is infrequently recorded in primary care records.[25] The influence of a partner on one’s care use may be different from that of a non-intimate co-resident although short-term co-residents are likely to be relatively rare among the over fifties. We did not consider the number or severity of conditions or acquisition of new conditions through follow-up, although our adjustment for age and socioeconomic deprivation may partly capture this. Although different lists of conditions were used to derive multimorbidity in the two datasets, there was considerable overlap and the main comparison of interest for this study was within-dataset differences between a multimorbid person co-resident with a healthy person and one co-resident with a multimorbid person. Finally, these findings should not be extrapolated to younger households, though previous work has highlighted the importance of the household health context for children’s healthcare use.[26]

### Implications

The largest cost differences between a multimorbid person co-resident with a healthy person and one co-resident with another multimorbid person were seen for community care and mental health care. However, most people did not have any activity for these services and consequently, confidence intervals were wide. It was not possible for us to extend this analysis to national data and this highlights the need for programmes to facilitate linkage of primary, secondary, community and social care.[27] Replication of the analysis for secondary care outcomes with a larger sample is also warranted, since the proportion with any hospital-based activity was also small. Our study indicates that CPRD is a suitable dataset for further exploration of the household health context and its impact on the health system and consequences for patient outcomes.

Our findings raise questions about how to deliver health and social care that acknowledges the household context for people with multimorbidity. For example, this could include scheduling community care to households or developing health care initiatives to households based on the principles of the group care approach.[28] If this is to be achieved then household context data will need to be made available to service providers within integrated care systems. This will require information governance standards to be upheld whilst at the same time ensuring household data can be shared for patient and public benefit.

## Conclusions

The number of people with multimorbidity is rising and our study suggests that multimorbidity may cluster in households with potential impacts on care systems, notably primary care and community care, and treatment burden for patients. In addition to preventive measures to modify risk factors that are common within households,[29] research is needed to test whether connecting service input across household members could lead to efficiency savings for health and care service providers or reduce treatment burden for those living with multimorbidity.

## Data Availability

We used two dataets for these analyses. 
A local sample was drawn from residents in Barking and Dagenham. De-identified data on their use of health and care services was available from linked data from local government services, health providers and health commissioners in the area. Information about accessing the data can be found here: https://www.carecity.london/your-blog/180-linking-datasets-for-better-population-health-management 
A national sample was drawn from the Clinical Practice Research Datalink (CPRD). Data access for this project has been approved (ISAC 17_150RMn2). Data used for this analysis is not publically available but anonymised patient datasets can be extracted for researchers against specific study specifications, following protocol approval from the Independent Scientific Advisory Committee (ISAC) https://www.cprd.com/

---

**Supplementary Table 1.**
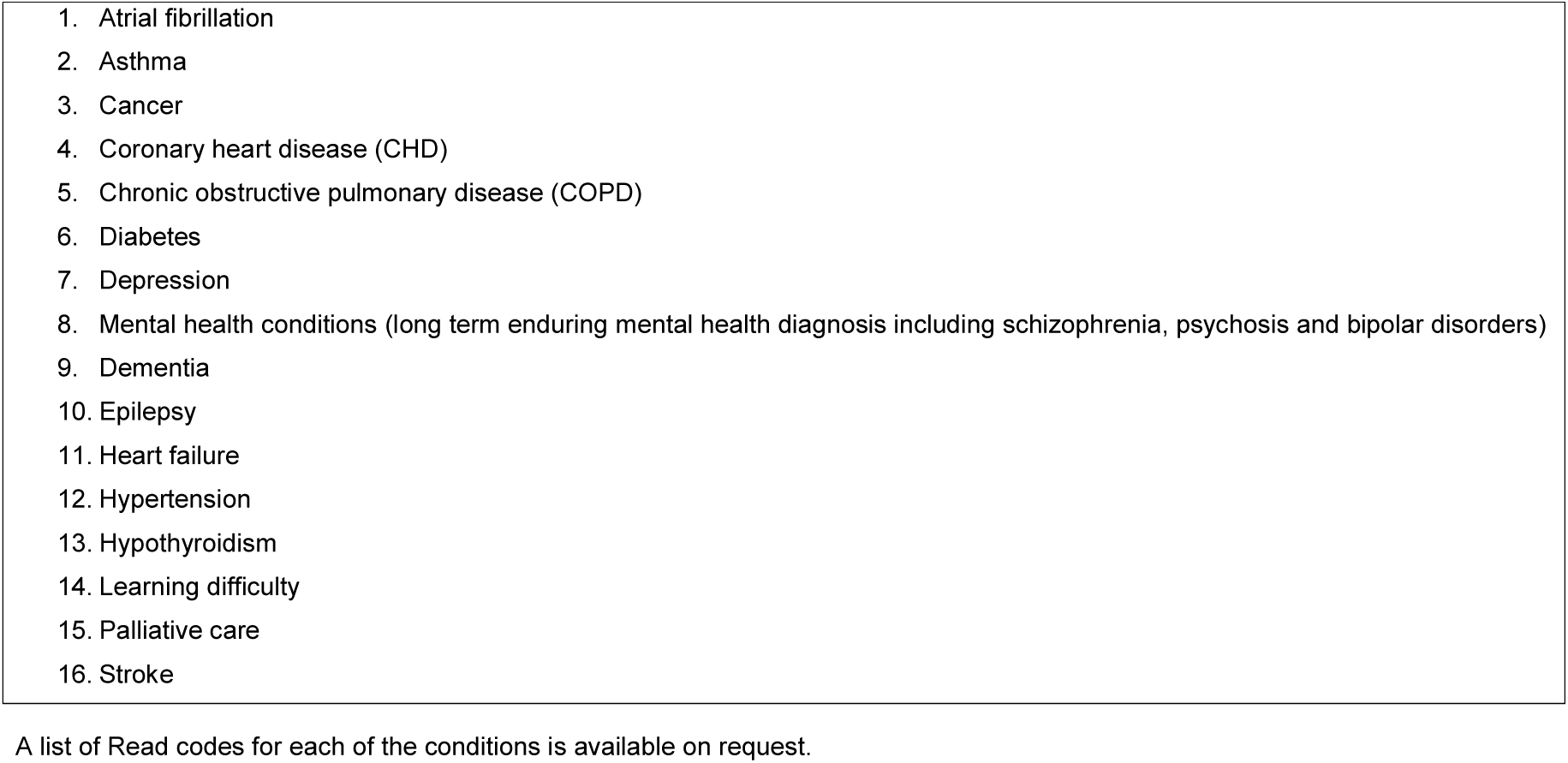
Long-term conditions used to define multimorbidity in Barking & Dagenham sample

**Supplementary Table 2.** Description of activity and cost of health and social care measures

|  | B & D sample | CPRD sample |
| --- | --- | --- |
| Primary care consultations | The activity data provided a count of GP and non-GP contacts per month, without a distinction between face-to-face consultations in the surgery, telephone consultations or home visits.<br>Unit costs were applied to GP and non-GP contacts using unit cost figures from the Personal Social Services Research Unit (PSSRU).<sup>28</sup> Unit costs included qualifications. For non-GP contacts, the hourly cost of a practice nurse was used. It was assumed that a nurse would see four people per hour. | Practice consultations, consultations involving a visit, and telephone consultations with a GP, nurse or other clinical staff were counted. [30]<br>Primary care costs were calculated by multiplying consultation duration by unit cost figures from the Personal Social Services Research Unit (PSSRU).[31] |
| Outpatient activity | The HRG national tariff for outpatient attendances was used. This varied according to the specialty the individual was visiting, whether it was a first appointment (new) or a follow-up and the commissioning threshold for new to follow-up ratios, which can lead to some appointments not having a reimbursement value. | Outpatient attendances with a valid HRG code were included. HRG4+ Reference Costs Groupers were used to identify the reference cost from the NHS Improvement Reference Costs for each year. |
| Admitted patient activity | Inpatient admissions and the corresponding HRG national tariff were used. Inpatient admissions were broken down into elective and non-elective.<br>Number of bed days was counted with day cases contributing 1 day. | Inpatient admissions with a valid HRG code were included for all main and up to 10 unbundled HRG codes. HRG4+ Reference Costs Groupers were used to identify the reference cost from the NHS Improvement Reference Costs for each year. Costs for inpatient admissions were broken down into elective; emergency; and all other admissions, which included day cases, maternity, and regular admissions.<br>Number of bed days was counted with day cases contributing 1 day. |
| Emergency department activity | The cost for each attendance varied depending on the type of emergency department (consultant-led emergency departments; consultant-led mono-specialty services; other types of minor injury departments; and NHS walk-in centres), whether the patient was admitted or not, and whether they arrived at the emergency department by | Emergency department attendances with a valid HRG code were included.<br>HRG4+ Reference Costs Groupers were used to identify the reference cost from the NHS Improvement Reference Costs for each year. |
|  | ambulance. These adjustments were made by the HRG grouper and the unit costs of each individual level activity reflected these adjustments. |  |
| Community and Mental health activity | Data from the patient level information and costing system from North East London NHS Foundation trust (the local provider of mental health and community services) were used to calculate unit costs for each component of activity. | N/A |
| Social care activity | Local authority social care costs were obtained from council data which lists the billed cost for each care package per week for each care recipient. This provided granularity on in-year changes to packages and resultant changes in costs. Data on self-funded social care was not available; neither was data on equipment, transport and home adaptation costs as these are held in different council departments. | N/A |

**Supplementary Table 3.**
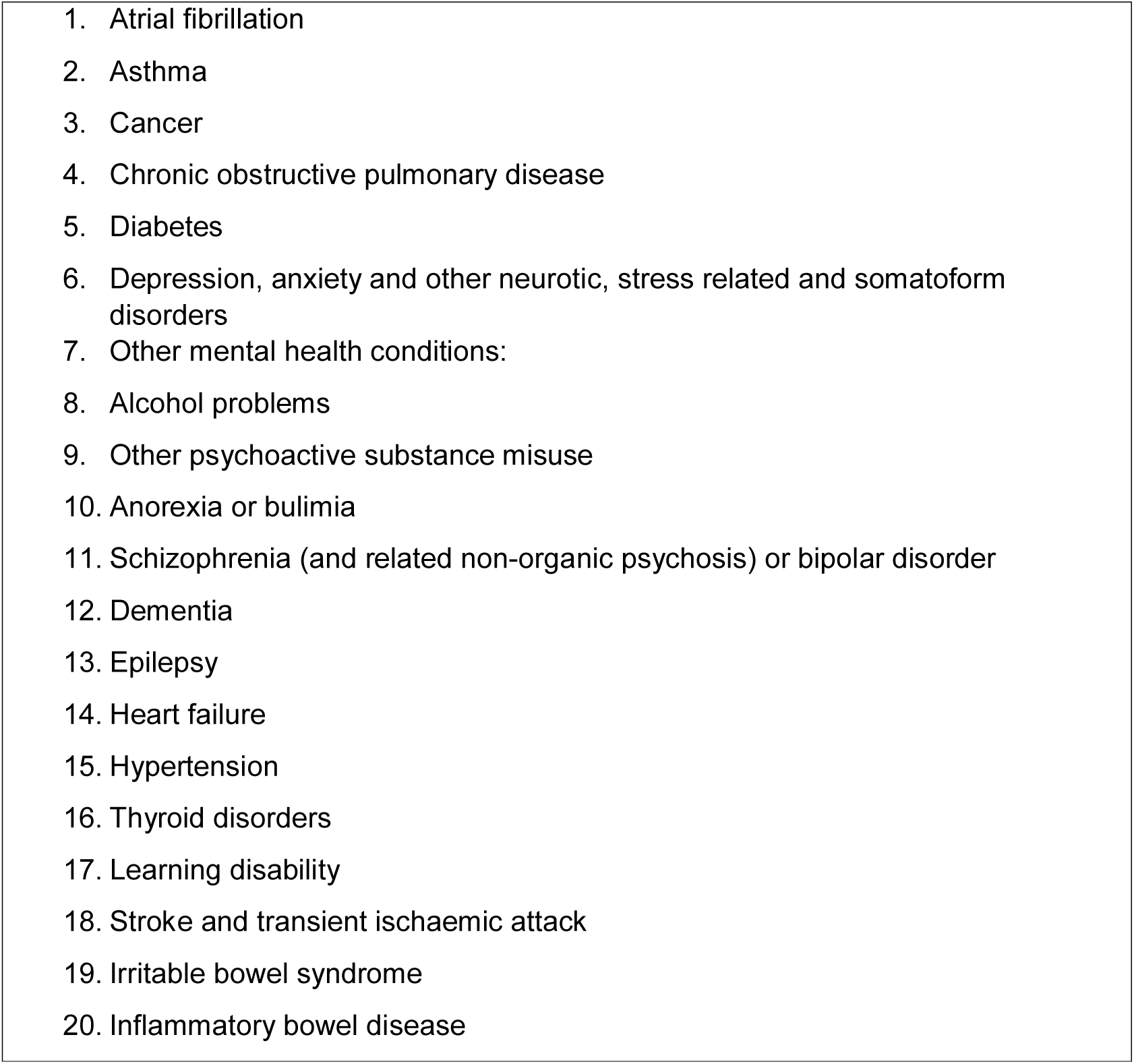
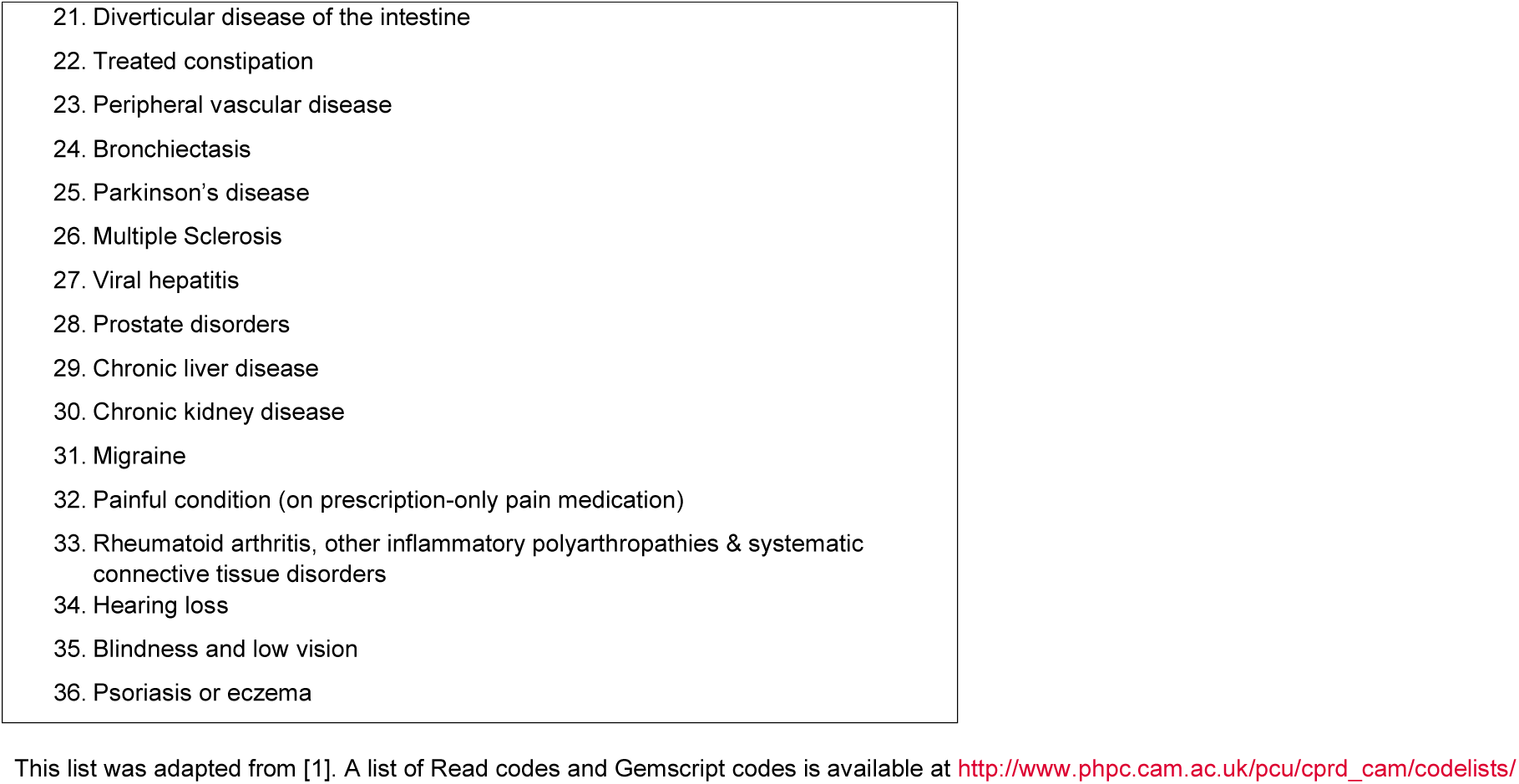
Long-term conditions used to define multimorbidity in CPRD sample

**Supplementary Table 4.** Description of health and social care activity and cost

|  | % with no<br>cost/activity | Mean activity<br>(number of<br>visits) | SD activity<br>(number of<br>visits) | Mean cost<br>(£) | SD cost<br>(£) |
| --- | --- | --- | --- | --- | --- |
| **B & D sample** |  |  |  |  |  |
| Primary care | 2.4 | 6.3 | 4.8 | 163.20 | 127.90 |
| Outpatient care | 37.5 | 3.2 | 4.5 | 233.20 | 351.60 |
| ED care | 65.4 | 0.3 | 0.9 | 64.10 | 159.40 |
| Non-elective admissions | 83.3 | 0.1 | 0.4 | 416.60 | 1525.20 |
| Elective admissions | 69.5 | 0.2 | 0.7 | 355.30 | 969.00 |
| Days in hospital | 82.3 | 1.1 | 4.7 | - | - |
| Community care | 76.6 | - | - | 297.8 | 1525.0 |
| Mental health services | 94.6 | - | - | 265.7 | 4212.5 |
| Local government funded social care | 96.4 | - | - | 263.7 | 2055.4 |
| **CPRD sample** |  |  |  |  |  |
| Primary care | 5.5 | 7.4 | 7.8 | 192.80 | 219.30 |
| Outpatient care | 39.1 | 2.6 | 4.9 | 360.30 | 851.50 |
| ED care | 73.0 | 0.2 | 0.6 | 56.10 | 166.30 |
| Non-elective admissions | 92.0 | 0.1 | 0.3 | 236.01 | 1075.19 |
| Elective admissions | 88.5 | 0.1 | 0.3 | 429.18 | 1365.38 |
| Days in hospital | 68.9 | 0.6 | 3.5 | - | - |

## References

[1] A. Cassell et al., “The epidemiology of multimorbidity in primary care: A retrospective cohort study,” Br. J. Gen. Pract., vol. 68, no. 669, pp. e245–e251, Apr. 2018.

[2] M. Stafford, A. Steventon, R. Thorlby, R. Fisher, C. Turton, and S. R. Deeny, “Understanding the health care needs of people with multiple health conditions,” London, 2018.

[3] H. Kasteridis P, Street A, Dolman M, Gallier L and W. I. k, Martin J, “The Importance of Multimorbidity in Explaining Utilisation and Costs Across Health and Social Care Settings: Evidence CHE Research Paper 96,” 2014.

[4] T. Lloyd, R. Brine, R. Pearson, M. Caunt, and A. Steventon, “Improvement Analytics Unit briefing Briefing: The impact of integrated care teams on hospital use in North East Hampshire and Farnham Consideration of findings from the Improvement Analytics Unit,” 2018.

[5] C. Sherlaw-Johnson, H. Crump, S. Arora, H. Holder, A. Davies, and R. Meaker, “Patient-centred care for older people with complex needs Evaluation of a new care model in outer east London,” 2018.

[6] HSCIC, “2010 Survey of Carers in Households 2009-10,” 2010.

[7] P. Hopman, M. J. Heins, J. C. Korevaar, M. Rijken, and F. G. Schellevis, “Health care utilization of patients with multiple chronic diseases in the Netherlands: Differences and underlying factors IZ,” Eur. J. Intern. Med., vol. 35, pp. 44–50, 2016.

[8] D. Meyler, J. P. Stimpson, and M. K. Peek, “Health concordance within couples: a systematic review.,” Soc. Sci. Med., vol. 64, no. 11, pp. 2297–2310, Jun. 2007.

[9] S. A. Patel et al., “Chronic disease concordance within Indian households: A crosssectional study.,” PLoS Med., vol. 14, no. 9, p. e1002395, Sep. 2017.

[10] P. Campbell, M. Shraim, K. P. Jordan, and K. M. Dunn, “In sickness and in health: A cross-sectional analysis of concordance for musculoskeletal pain in 13,507 couples.,” Eur. J. Pain, vol. 20, no. 3, pp. 438–446, Mar. 2016.

[11] K. Dreyer, A. Steventon, R. Fisher, and S. R. Deeny, “The association between living alone and health care utilisation in older adults: A retrospective cohort study of electronic health records from a London general practice,” BMC Geriatr., vol. 18, no. 1, Dec. 2018.

[12] G. Payne, A. Laporte, R. Deber, and P. C. Coyte, “Counting backward to health care’s future: Using time-to-death modeling to identify changes in end-of-life morbidity and the impact of aging on health care expenditures,” Milbank Q., vol. 85, no. 2, pp. 213– 257, 2007.

[13] K. Barnett, S. W. Mercer, M. Norbury, G. Watt, S. Wyke, and B. Guthrie, “Epidemiology of multimorbidity and implications for health care, research, and medical education: a cross-sectional study.,” Lancet (London, England), vol. 380, no. 9836, pp. 37–43, Jul. 2012.

[14] Rupert A Payne et al., “Development and validation of the Cambridge Multimorbidity Score,” C. | Febr., vol. 3, pp. 107–121, 2020.

[15] E. Herrett et al., “Data Resource Profile: Clinical Practice Research Datalink (CPRD),” Int. J. Epidemiol., vol. 44, no. 3, pp. 827–836, Jun. 2015.

[16] U. of C. Department of Public Health and Primary Care, “CPRD@Cambridge - Code lists.” [Online]. Available: http://www.phpc.cam.ac.uk/pcu/cprd_cam/codelists/. [Accessed: 29-Jan-2020].

[17] B. Babitsch, D. Gohl, and T. Von Lengerke, “Re-revisiting Andersen’s Behavioral Model of Health Services Use: a systematic review of studies from 1998-2011 Das Verhaltensmodell der Inanspruchnahme gesundheitsbezogener the retrieved articles for possible inclusion using a three-step selection process (1. title/author, 2. abstract, 3. full text) with pre-defined inclusion OPEN ACCESS,” 2012.

[18] C. H. Van Houtven and E. C. Norton, “Informal care and health care use of older adults,” J. Health Econ., vol. 23, no. 6, pp. 1159–1180, Nov. 2004.

[19] M. Wagner and M. Brandt, “Long-term Care Provision and the Well-Being of Spousal Caregivers: An Analysis of 138 European Regions,” Journals Gerontol. - Ser. B Psychol. Sci. Soc. Sci., vol. 73, no. 4, pp. e24–e34, Apr. 2018.

[20] G. P. A. Thomas, C. L. Saunders, M. O. Roland, and C. A. M. Paddison, “Informal carers’ health-related quality of life and patient experience in primary care: Evidence from 195,364 carers in England responding to a national survey,” BMC Fam. Pract., vol. 16, no. 1, May 2015.

[21] Y. Tommis, C. A. Robinson, D. Seddon, B. Woods, J. Perry, and I. T. Russell, “Carers with chronic conditions: Changes over time in their physical health,” Chronic Illn., vol. 5, no. 3, pp. 155–164, 2009.

[22] “LC3301EW Provision of unpaid care by general health by sex by age.” [Online]. Available: https://www.nomisweb.co.uk/census/2011/lc3301ew.

[23] “The NHS Long Term Plan,” 2019.

[24] Department of Health and Social Care, “Carers Action Plan 2018 to 2020: Supporting carers today,” 2018.

[25] A. Jain, A. J. Van Hoek, J. L. Walker, R. Mathur, L. Smeeth, and S. L. Thomas, “Identifying social factors amongst older individuals in linked electronic health records: An assessment in a population based study,” PLoS One, vol. 12, no. 11, Nov. 2017.

[26] K. Dreyer, R. A. P. Williamson, D. S. Hargreaves, R. Rosen, and S. R. Deeny, “Associations between parental mental health and other family factors and healthcare utilisation among children and young people: a retrospective, cross-sectional study of linked healthcare data,” BMJ Paediatr. Open, vol. 2, no. 1, p. e000266. Jul. 2018.

[27] “Local Health and Care Record Exemplars A summary OFFICIAL 2 Local Health and Care Record Exemplars.”

[28] W. J. J. Edelman D, McDuffie JR, Oddone E, Gierisch JM, Nagi A, “Shared medical appointments for chronic medical conditions: a systematic review. VAESP Project #09-010,” 2012.

[29] Academy of Medical Sciences, Multiple morbidities as a global health challenge. London: Academy of Medical Sciences, 2015.

[30] K. Dreyer, W. Parry, W. Jayatunga, and S. Deeny, “A descriptive analysis of health care use by high- cost, high-need patients in England,” 2019.

[31] L. Curtis and A. Burns, “Unit Costs of Health and Social Care 2015.,” 2015.

